# Determinants of Cerebrovascular Reserve in Patients with Significant Carotid Stenosis

**DOI:** 10.1101/2020.11.15.20232157

**Authors:** Joseph P Archie

## Abstract

**Introduction:** In patients with 70% to 99% diameter carotid artery stenosis cerebral blood flow reserve may be protective of future ischemic cerebral events. Reserve cerebral blood flow is created by brain auto-regulation. Both cerebral blood flow reserve and cerebrovascular reactivity can be measured non-invasively. However, the factors and variables that determine the availability and magnitude and of reserve blood flow remain poorly understood. The availability of reserve cerebral blood flow is a predictor of stroke risk. The aim of this study is to employ a hemodynamic model to predict the variables and functional relationships that determine cerebral blood flow reserve in patients with significant carotid stenosis.

**Methods:** A basic one-dimensional, three-unit (carotid, collateral and brain) energy conservation fluid mechanics blood flow model is employed. It has two distinct but adjacent blood flow components with normal cerebral blood flow at the interface. In the brain auto-regulated blood flow component cerebral blood flow is maintained normal by reserve flow. In the brain pressure dependent blood flow component cerebral blood flow is below normal because cerebral perfusion pressure is below the lower threshold value for auto-regulation. Patient specific values of collateral vascular resistance are determined from a model solution using clinically measured systemic and carotid arterial stump pressures. Collateral vascular resistance curves illustrate the model solutions for reserve and actual cerebral blood flow as a function of percent diameter carotid artery stenosis and mean systemic arterial pressure. The threshold cerebral perfusion pressure value for auto-regulation is assumed to be 50 mmHg. Normal auto-regulated regional cerebral blood flow is assumed to be 50 ml/min/100g. Cerebral blood flow and reserve blood flow solutions are given for systemic arterial pressures of 80, 90, 100, 110 and 120 mmHg and for three patient specific collateral vascular resistance values, Rw = 1.0 (mean patient value), Rw = 0.5 (lower 1 SD) and Rd = 3.0 (upper 1 SD).

**Results:** Reserve cerebral blood flow is only available when a patients cerebral perfusion pressure is in the normal auto-regulatory range. Both actual and reserve cerebral blood flows are primarily from the carotid circulation when carotid stenosis is less than 60% diameter. Between 60% and 75% stenosis the remaining carotid blood flow reserve is utilized and at higher degrees of stenosis all reserve flow is from the collateral circulation. The primary independent variables that determine actual and reserve cerebral blood flow are mean systemic arterial pressure, degree of carotid stenosis and patient specific collateral vascular resistance. Approximate 16% of patients have collateral vascular resistance greater than 5.0 and are predicted to be at high risk of cerebral ischemia or infarction with progression to severe carotid stenosis or occlusion. The approximate 50% of patients with a collateral vascular resistance less than 1.0 are predicted to have adequate cerebral blood flow with progression to carotid occlusion, and most maintain some reserve. Clinically measured values of cerebral blood flow reserve or cerebrovascular reactivity are predicted to be unreliable without consideration of systemic arterial pressure and degree of carotid stenosis. Reserve cerebral blood flow values measured in patients with only moderate 60% to 70% carotid stenosis are in general too high and variable to be of clinical value, but are most reliable when measured near 80% diameter stenosis and considered as percent of the maximum reserve blood flow. Patient specific measured reserve blood flow values can be inserted into the model to calculate the collateral vascular resistance.

**Conclusions:** Predicting cerebral blood flow reserve in patients with significant carotid stenosis is complex and multifactorial. A simple cerebrovascular model predicts that patient specific collateral vascular resistance is an excellent predictor of reserve cerebral blood flow in patients with significant carotid stenosis. Cerebral blood flow reserve measurements are of limited value without accounting for systemic pressure and actual percent carotid stenosis. Asymptomatic patients with severe carotid artery stenosis and a collateral vascular resistance greater than 1.0 are at increased risk of cerebral ischemia and may benefit from carotid endarterectomy.

## Introduction

Optimal management strategy for patients with asymptomatic moderate to severe atherosclerotic carotid artery stenosis remains challenging because the value of carotid surgery for preventing stroke is yet to be determined. Even with best current medical management including statins and anti-platelets drugs, the risk of ischemic stroke from emboli or inadequate cerebral blood flow remains a major concern. Independent of the outcomes of ongoing prospective randomized trials of carotid surgery versus best medical treatment there is likely to be a sub-set of asymptomatic patients with significant carotid stenosis that would benefit from carotid endarterectomy to prevent stroke. Potential inclusive criteria are patients with unstable carotid plaque or inadequate reserve cerebral blood flow. While carotid plaque stability-embologeneity, degree of carotid stenosis and estimates of cerebrovascular blood flow reserve have been extensively studied, there remains no scientifically acceptable criteria for recommending reconstructive carotid surgery in these patients. A recent review and meta analysis (1, 2) of studies reporting cerebrovascular reserve blood flow or cerebrovascular reactivity in patients with greater than 70% carotid artery stenosis found on that those with impaired reserve or reactivity had a significantly higher stroke/TIA rate than those with measurable reserve/reactivity deemed adequate over a three year follow-up. This was independent of carotid stenosis or occlusion, with or without symptoms and method of measuring reserve and reactivity. Potential key determinants of cerebrovascular blood flow reserve not considered include the degree of carotid stenosis, systemic arterial blood pressure and a measure of the availability of collateral blood flow primarily via the circle of Willis. While it has been suggested that several of these independent variables may be involved in determining cerebrovascular blood flow reserve, the evidence for this is meager, conflicting and confusing. Over a half-century ago Lassen (3) found auto-regulation of cerebral blood flow in man to be approximately 50ml/min/100g over a range of mean threshold cerebral perfusion pressures of 50mmHg up to 160mmHg. This fundamental hemodynamic relationship is the basis for a simple hemodynamic model that predicts cerebral blood flow and reserve blood flow in patients with carotid stenosis (4). The aim of this study is to utilize a hemodynamic model to predict the variables and functional relationships that determine cerebral blood flow reserve in patients with significant carotid stenosis.

## Methods

### The Model

A classical one-dimensional Bernoulli fluid mechanics energy conservation model is used for each of the three anatomically separate but arterially connected units: carotid, collateral and brain. The hemodynamics for each unit is pressure gradient (P) equals vascular resistance (R) times volumetric blood flow (Q), or P = RQ. Both pressure and blood flow are measurable variables, while vascular resistance is a derived and calculated variable defined as R = P/Q. It can be a constant or a variable. The one-dimensional lumped parameter model is based on mean pressure and mean flow. The carotid and collateral components are in parallel with input mean arterial pressure Pa and output cerebral perfusion pressure Pp to the brain in series (4). The algebraic equation is; Pa = (Rt + Rb)Qn, where Pa is mean systemic arterial pressure and Rt is the parallel carotid and collateral vascular resistances Rc and Rw and Rt = RcRw/(Rc + Rw) or 1/Rt = 1/Rc + 1/Rw. Brain vascular resistance is Rb and Qn is normal cerebral blood flow. The three component vascular resistances, carotid (Rc), collateral (Rw) and brain (Rb), are unique independent variables. The component equations are; (Pa – Pp) = Rt Qn and Pp = RbQn, with (Pa – Pp) = RcQc = RwQw where Qn = Qc + Qw. Carotid resistance is assumed to equal percent area stenosis, Rc = 1/[(1/X^2^) – 1], where X is fractional percent diameter stenosis. When there is no carotid stenosis X = 0 and Rc = 0. When Rc = 1.0, X = 0.707 or 70.7% diameter stenosis. With carotid occlusion Rc = ∞, 100% carotid stenosis, there is no carotid blood flow. Collateral vascular resistance Rw is assumed to be due to a patient’s circle of Willis. The lower cerebral perfusion pressure threshold for auto-regulation, 50mmHg, dictates that the model has two interfaced components. The upper auto-regulation zone where Q is the sum of normal cerebral blood flow, Qn and the reserve flow Qr, or Q = Qn + Qr and Rb = Q/Qn, a variable. The interface is normal cerebral blood flow Qn. In the lower linear perfusion pressure dependent zone cerebral blood flow Q is less than normal (Qn) and equals perfusion pressure, Q = Pp, Rb = 1.0 = Pp lower threshold/Qn = 50/50 = 1.0, a linear relationship. Powers recognized the clinical importance of this two-component model three decades ago (5). When the carotid is occluded (Rc = ∞), Rt = Rw and cerebral perfusion pressure Pp is the carotid stump pressure Ps. The solution to this model equation is Rw = Rb (Pa/Ps – 1). This is a powerful tool because Rw is patient specific and defines a patient’s collateral cerebral blood flow potential with a single number - value (4). Further, Rw patient specificity allows model solutions for reserve and actual cerebral blood flow versus percent diameter carotid stenosis and mean systemic arterial pressure to be presented graphically by Rw curves. When carotid stump pressure Ps is equal to or less than the lower threshold perfusion pressure for cerebrovascular regulation, Pp = 50 mmHg, cerebral blood flow is pressure dependent and Rw = 1.0 and Rw = (Pa/Ps – 1). When Ps is greater than 50mmHg threshold the solution is Rw = (Ps/50) (Pa/Ps -1).

### Solutions For Cerebrovascular Reserve Using Clinical Patient Specific Collateral Vascular Resistance

The physiologic necessity for the model to have two distinct components, auto-regulation and pressure dependent, also dictates that all reserve blood flow is in the auto-regulatory zone. Further, collateral vascular resistance, Rw, is the unifying variable that allows solutions of the complete model. The cerebral blood flow, Q, solutions given in the results seven figures illustrate the dependence of reserve flow on arterial pressure Pa, percent diameter carotid stenosis. Collateral vascular resistance, Rw, is the unifying index that predicts reserve blood flow.

### Determining Collateral Vascular Resistance with Patient Measured Reserve Blood Flow

Direct measurement of collateral vascular resistance, Rw, in patients requires both mean systemic arterial pressure, Pa and carotid stump pressure, Ps. Obtaining the latter can currently only be done by invasively measuring directly at surgery or with balloon catheter temporary carotid occlusion. The model solution for collateral vascular resistance offers an alternative non-invasive method; Rw = Rc (Pa/Qn – Rb)/[Rc – (Pa/Qn – Rb)]. The required measurements are % diameter carotid stenosis, (X/100, to calculate Rc), mean systemic arterial pressure, Pa, and cerebral blood flow at maximum vasodilation, Q. In the auto regulation range Rb = Q/Qn where Qn is 50ml/min/100g and either Q or Q/Qn is measured. Carotid vascular resistance is Rc = 1/[(1/X^2^) – 1]. The difficult part is accurate reserve blood flow measurements. Both qualitative and quantitative estimates of Rw are given using published data. A quick way to calculate Rw is to find Rt = (Pa/Qn - Q/Qn), find Rc = 1/[(1/X^2^) – 1] and calculate Rw = 1/(1/Rt – 1/Rc).

## RESULTS

Cerebrovascular Reserve Predicted By Collateral Vascular Resistance Two key findings results from a previous study of these cerebrovascular model equations are the foundation of these results (4). First, collateral vascular resistance can be determined from systemic arterial pressure and carotid stump pressure, Rw = Rb(Ps/Pa – 1), and second, Rw is patient specific over the normal range of systemic arterial pressures. Based on measurements of Ps and Pa in 1,360 patients with significant carotid stenosis (6) the mean value of Rw is approximately 1.0 and the standard deviations are approximately Rw = 3.0 and Rw = 0.5. This means that 68%, about two thirds, of these patients had Rw values between 0.5 and 3.0. The general conclusion of studies reporting measured cerebrovascular reserve/reactivity in patients with greater than 70% carotid diameter stenosis is that the availability collateral blood flow is the key to preventing future cerebral ischemia (7, 8). However, quantitation of the amount of reserve cerebral blood flow is rare and the impact of systemic arterial pressure and precise degree of carotid stenosis on reserve values is not considered. Figures 1, 2, 3 and 4 illustrate the effect of systemic arterial pressure, Pa, and degree of diameter carotid stenosis, X, on reserve cerebral blood flow at four patient specific Rw values. Figure 1 is when Rw = ∞, no collateral blood flow, a theoretical value because an accurately measured carotid stump pressure of zero-mmHg has not been reported. Figure 1 is the model solution in the absence of collateral flow. The transition from normal carotid blood flow and reserve blood flow from the auto-regulation model component to the pressure dependent component where cerebral perfusion pressure is less than 50 mmHg occurs from 61% to 76% stenosis as systemic arterial pressure increases from 80mmHg to 120mmHg. This means that there is no contribution from the carotid artery to cerebrovascular reserve at higher degrees of stenosis. At moderate to severe carotid stenosis essentially all reserve cerebral blood flow must be collateral in origin. Figures 2, 3 and 4 when Rw = 3.0, 1.0 and 0.5 respectively, the spectrum of 68% of patient’s Rw values, illustrate the progressive improvement in reserve cerebral blood flow as collateral vascular resistances decreases from infinity (Rw = ∞) to near zero (Rw = 0). Most patients with a collateral vascular resistance Rw less than 1.0 are predicted to have adequate collateral reserve after 70% stenosis. The average patient (Rw ∼1.0) is predicted to have reserve cerebral blood flow up to 90% stenosis and only a slightly below normal with occlusion. This is not the case with the higher degrees of collateral vascular resistance. For example, at Rw = 3.0 and greater values about 23% of patients are predicted to have inadequate reserve cerebral blood flow to prevent a decrease in normal cerebral flow, ischemic symptoms or stroke. These results clearly indicate that systemic arterial pressure is a major determinant of cerebral reserve flow and must be integrated into clinical assessment of cerebrovascular reserve. Further, an accurate measure of degree of carotid stenosis is mandatory. The progression of predicted values of reserve blood flow between 70% and 90% diameter stenosis is profound and relatively independent of systemic pressure.

**Figure 1.**
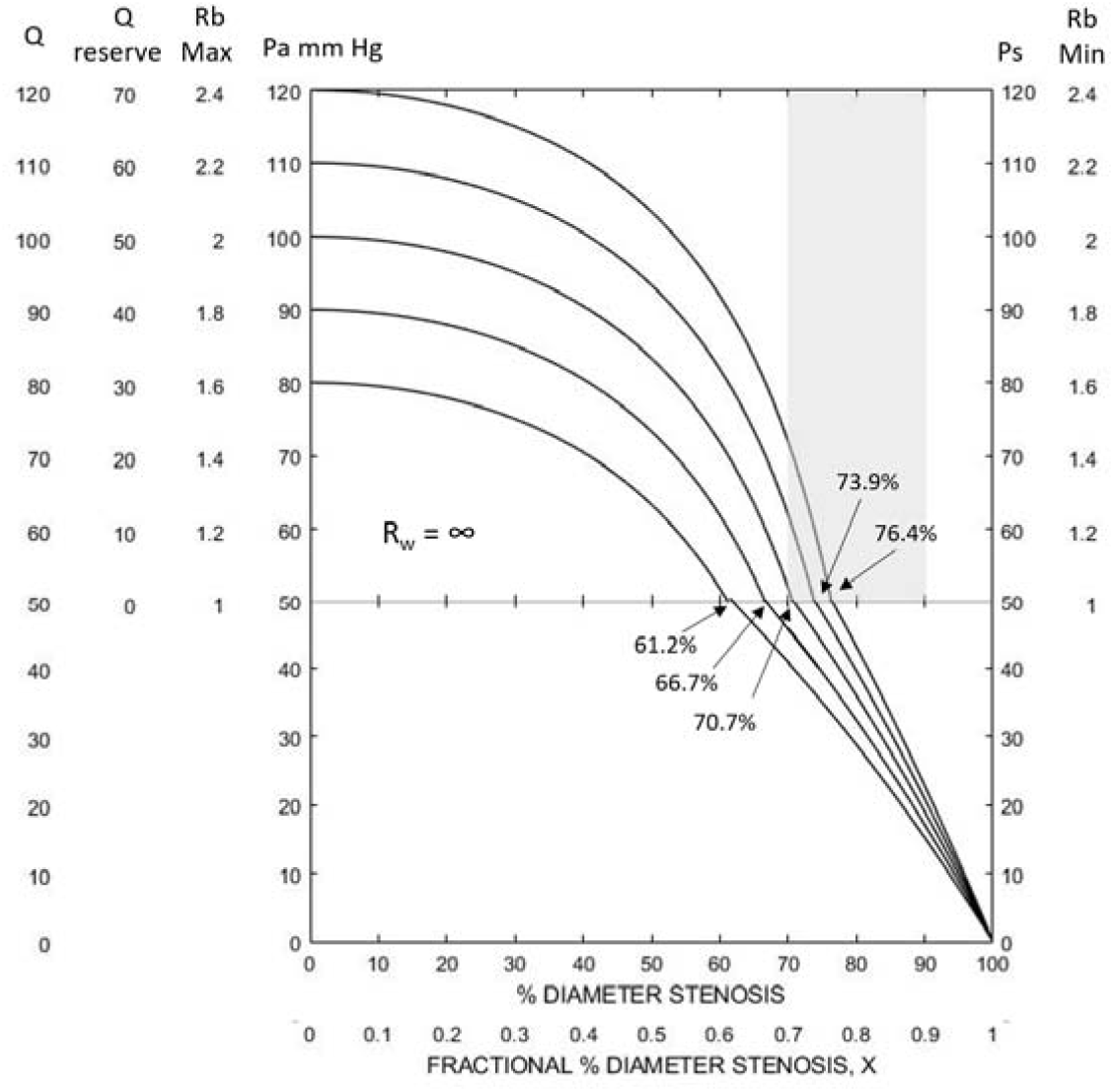
Reserve and actual cerebral blood flow curves for patient specific collateral vascular resistance Rw = ∞ (no collateral blood flow) over a range of mean systemic arterial pressures. Actual normal cerebral blood flow of 50ml/min/100g is at the interface of the auto-regulation and pressure dependent components (Q = 50) except at high percent stenosis when Rw curve falls into the pressure dependent cerebral ischemic zone. The collateral vascular resistance is infinity, Rw = ∞, because carotid stump pressure is zero, Ps = 0. These are theoretical values never measured. With stenosis progression to about 60% diameter the carotid artery provides reserve and actual cerebral blood flow. With carotid stenosis progression reserve flow becomes depleted between 61% and 76% stenosis depending on systemic arterial pressure. In the absence of collateral circulation, 80% stenosis produces significant cerebral ischemia and stroke is predicted at 90% stenosis. While this is a theoretical worst-case scenario, patients with poor collateral blood flow as determined by high collateral vascular resistance values have a significant risk of stroke with carotid stenosis progression as illustrated in Figure 2.

**Figure 2.**
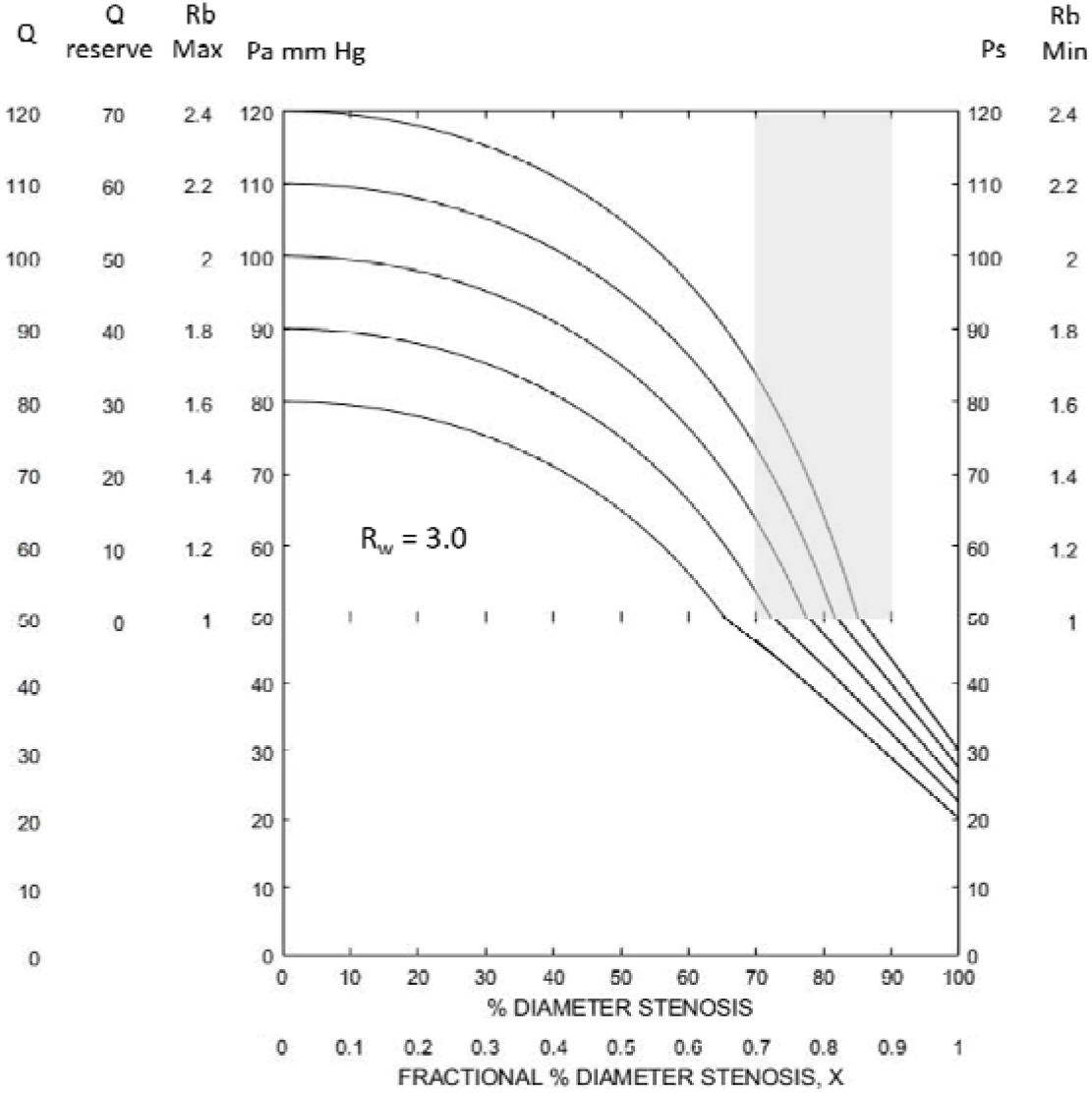
Reserve and actual cerebral blood flow curves for patient specific collateral vascular resistance Rw = 3.0, the upper one standard deviation from the normal Rw value of 1.0. Depending on systemic arterial pressure most of these patients are predicted to have some blood flow reserve at 70% stenosis but after 85% all develop some degree of cerebral ischemia. With carotid occlusion all patients are predicted to have cerebral perfusion pressure below 30mmHg, the threshold for cerebral ischemic symptoms. Clearly, patients with Rw > 3.0 should be considered for carotid endarterectomy.

**Figure 3.**
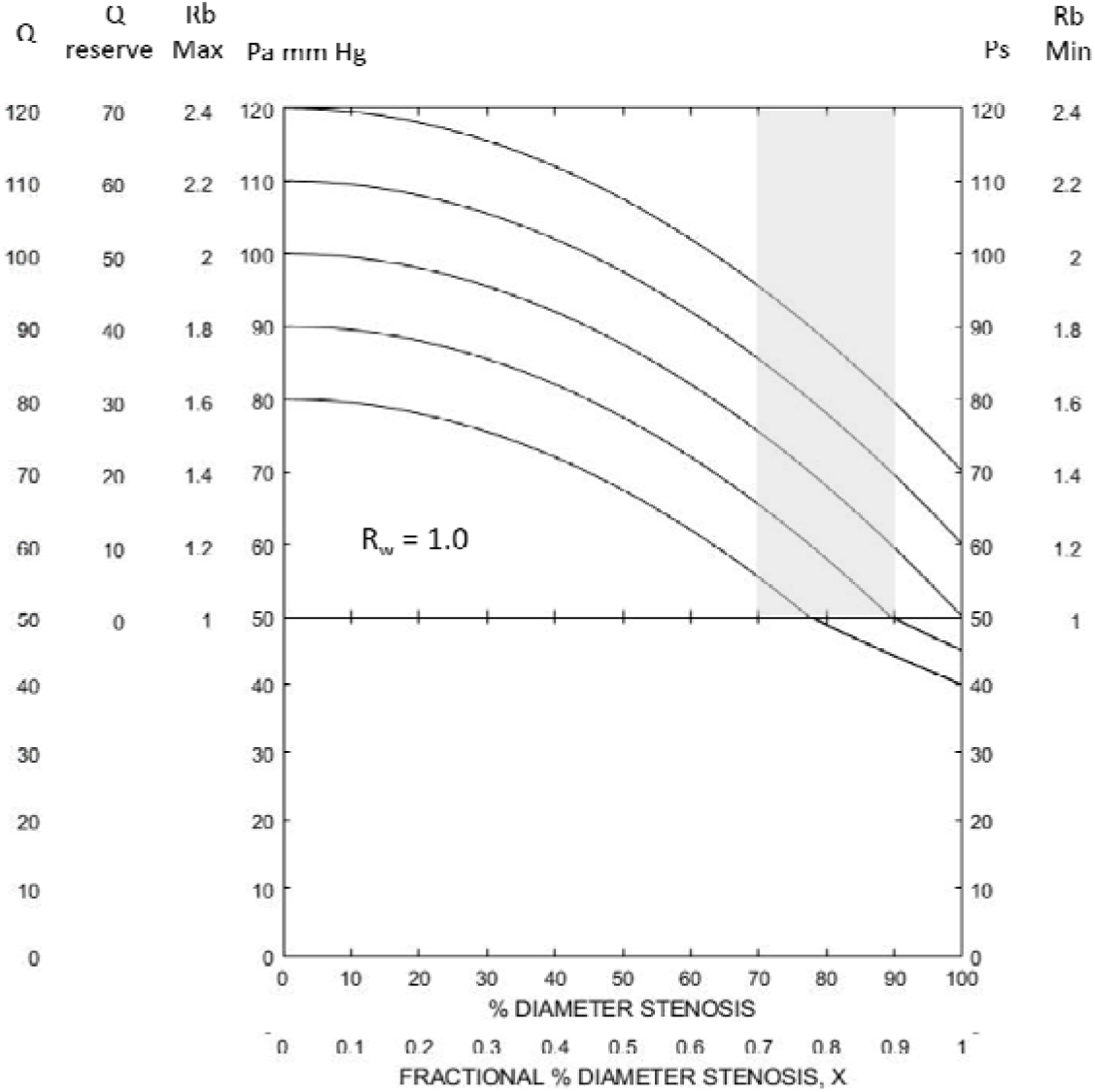
These Rw = 1.0 curves at different mean systemic pressures represent the average patient with significant carotid stenosis. While mild hypertension shifts the curves to the right improving reserve blood flow and the possibility of mild regional cerebral ischemia. Patients with a hemisphere Rw value near 1.0 are in general protected from ischemic hemodynamic stroke with carotid occlusion.

**Figure 4.**
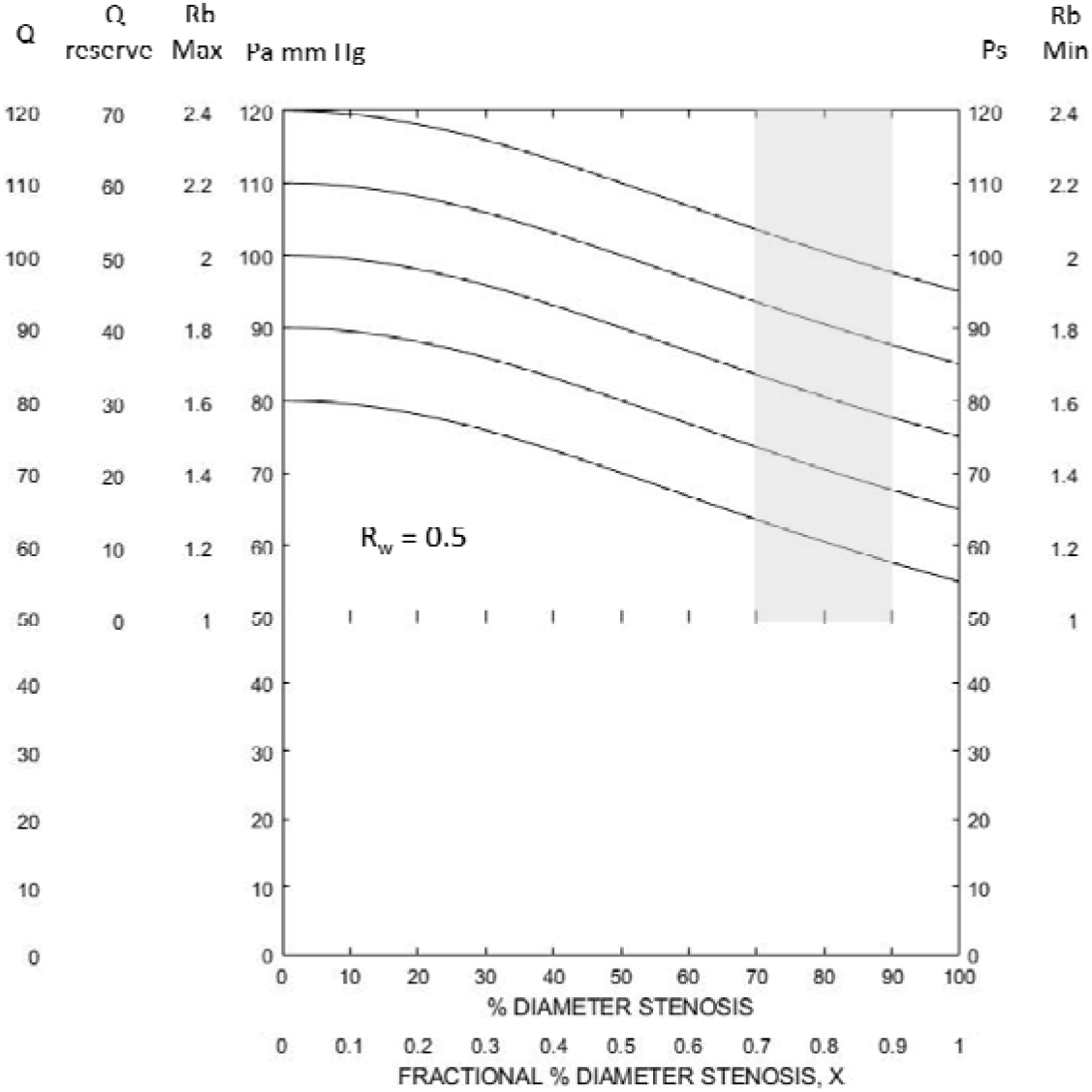
These Rw = 0.5 patients are in the safety zone with excellent reserve blood flow even at low normal systemic pressure and high-grade carotid stenosis. This group and perhaps all patients with Rw < 1.0, approximately half of all patients with significant carotid stenosis, are predicted to not benefit from carotid endarterectomy and should not be subjected to the perioperative risk.

While the impact of arterial pressure and degree of stenosis on blood flow reserve is significant, the primary determinant is collateral vascular resistance. This is illustrated in Figures 5 and 6 with patient’s systemic arterial pressure of 90 mmHg and 100 mmHg, perhaps in the normal window. The Rw = ∞ curve is included to illustrate the absence of a carotid contribution to reserve. Half of patients are predicted to maintain normal cerebral blood flow at carotid occlusion and most have adequate reserve at high-grade stenosis. The majority of patients are predicted to have adequate reserve cerebral blood flow at 80% stenosis.

**Figure 5.**
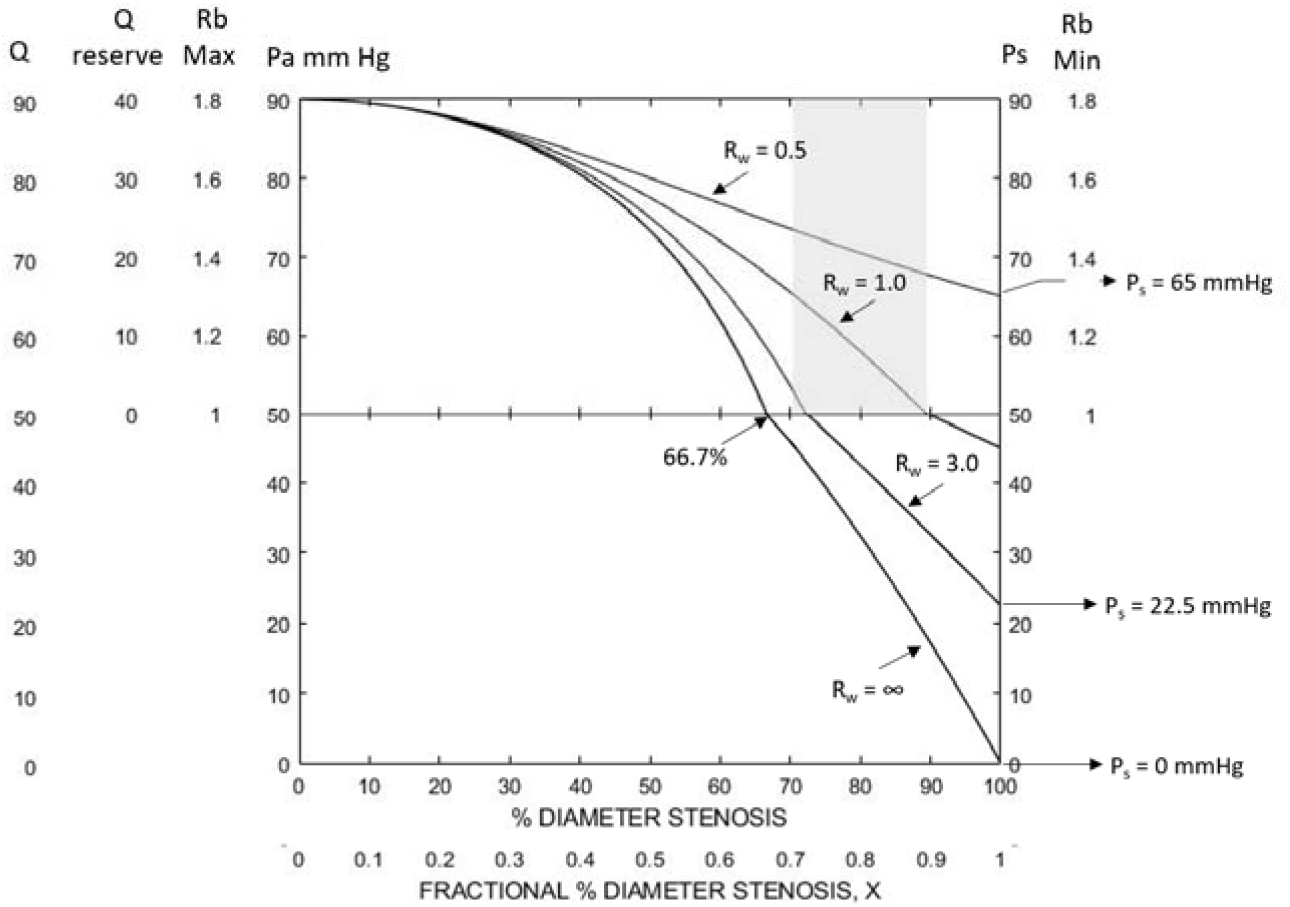
These are the Rw curves for a spectrum of patient’s Rw values at a mean systemic arterial pressure of 90mmHg. The Rw = ∞ curve is included illustrate that carotid blood flow is predicted to provide both reserve and normal cerebral blood flow up to about 67% stenosis after which collateral blood flow must be adequate to prevent cerebral infarction. Patients with Rw values greater than 1.0 are at risk of inadequate cerebral blood flow at high-grade stenosis or occlusion.

**Figure 6.**
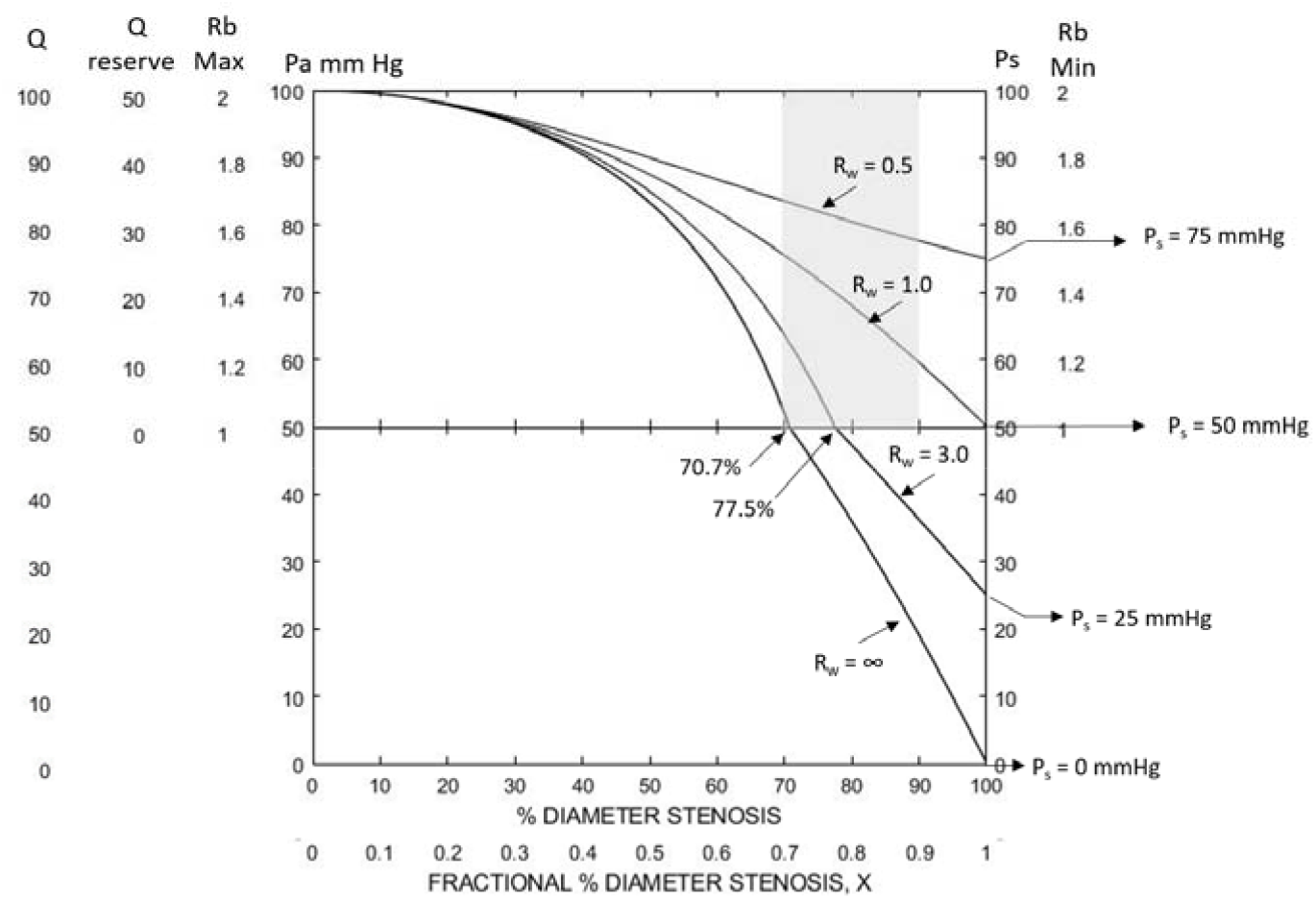
This is identical to Figure 5 except that mean systemic blood pressure is 100mmHg. This slightly higher systemic arterial pressure improves reserve and maintains normal cerebral blood flow, when Rw = 1.0.

The cerebral blood flow threshold for irreversible ischemia of 18ml/min/100g was first measured by Boysen (9). This worse case scenario is given in Figure 7 for Pa values 80mmHg to 120mmHg. As expected, the Rw values are high, 3.4 to 5.6, and these patients should clearly not be considered for carotid surgery.

**Figure 7.**
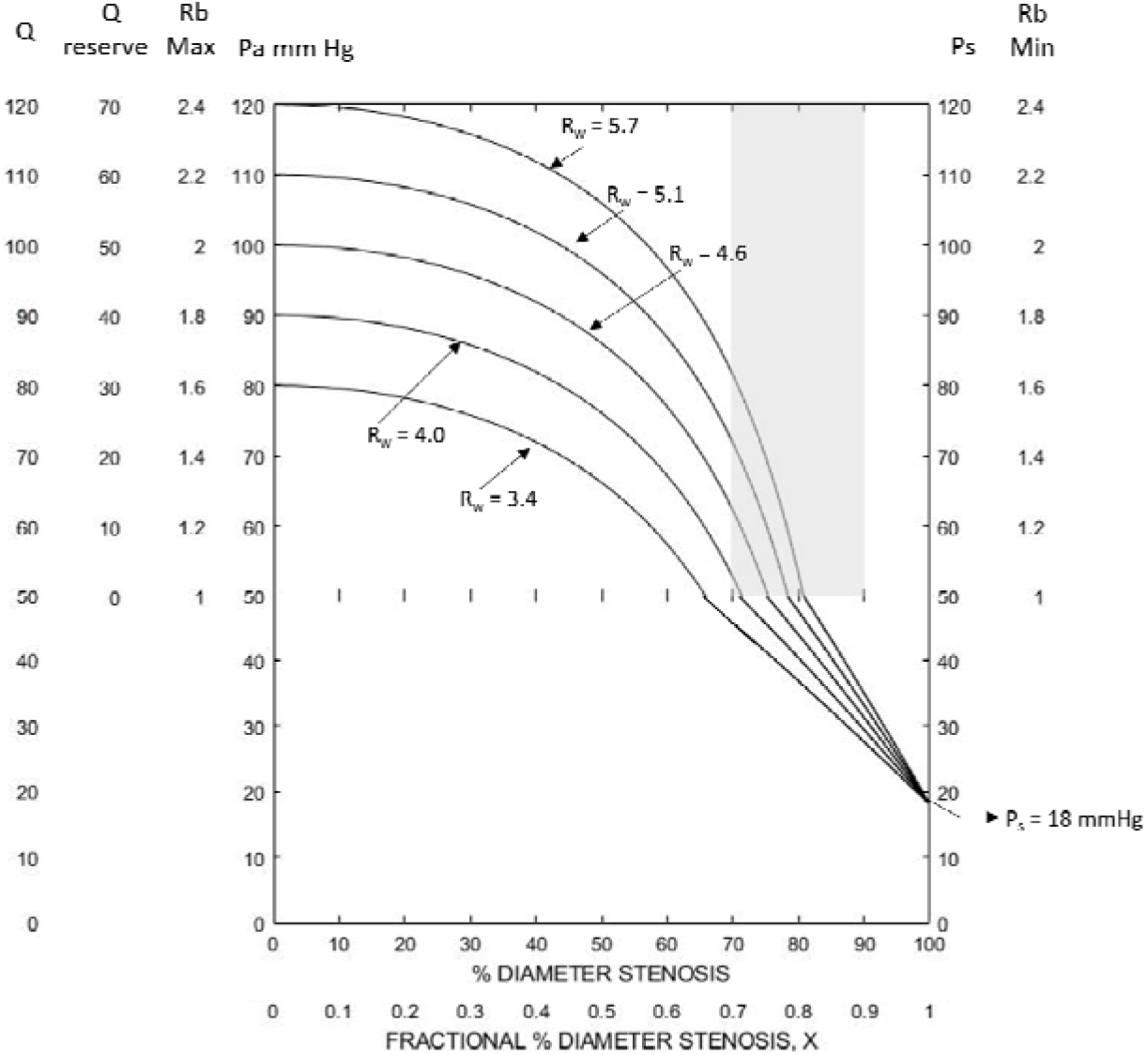
This illustrates the false security of assuming that adequate reserve blood flow at 60% to 70% stenosis predicts adequate collateral flow.

### Collateral Cerebral Vascular Resistance Using Cerebrovascular Reserve Blood Flow Measurements

The results g in Figures 1 to 4 illustrate that the amount of cerebral blood flow

reserve predicted by collateral vascular resistance, Rw, is highly dependent on mean systemic arterial pressure, Pa, and degree of carotid stenosis, X. Collateral vascular resistance, Rw, can be calculated from clinically measured cerebral vascular reserve blood flow, Qr, normal cerebral blood flow, Qn, Pa and X. As given in methods, Rw is calculated from the algebraic equations; Rw = 1/(1/Rt – 1/Rc) where Rc = 1/[(1/X^2^) – 1], Rt = (Pa/Qn - Q/Qn) and Q = Qr + Qn.

Several techniques have been used to measure or estimate cerebral blood flow reserve-reactivity in patients with significant carotid stenosis with the aim of stroke risk stratification. The dynamic method measures the response to a rapid change in systemic arterial pressure as with deflation of thigh blood pressure cuffs. The static uses pharmacologic maximal cerebral vasodilation with acetazolamide or CO2.

Reserve-reactivity is measured directly by regional volumetric cerebral blood flow with CT, MRI, PET or SPECT scans or indirectly by middle cerebral artery mean blood velocity with trans cranial Doppler. While two methods correlate they give different numerical results when used with acetazolamide vasodilation (10). Most published cerebral blood flow reserve or cerebrovascular reactivity results must be considered qualitatively as either impaired or adequate due to the absence of accepted criteria or values. This is easy to appreciate when considering the array of predicted reserve blood flow values arrived at without consideration of the effect of systemic arterial pressure of accurate degree of stenosis. These concerns are expressed in the review and meta-analysis of Gupta (2). Of the 13 studies that met the inclusion criteria of greater than 70% diameter carotid stenosis and at least one-year follow-up for stroke or TIA, six had patients only carotid occlusion and four with only carotid stenosis. From a table, 24/233 patients with only carotid occlusion and adequate reserve/reactivity had a subsequent stroke (10%) versus 54/169 (32%) of patients with impaired reserve/reactivity (P < 0.001). For patients with only carotid stenosis this was 7/166 (4.2%) stroke/TIA and adequate reserve versus 24/102 (24%) with impaired reserve (P <0.001). Their meta-analysis confirmed the significant positive relationship between cerebrovascular reserve/reactivity impairment and development of stroke. Missing from this analysis and the majority of publications is consideration of systemic arterial pressure or of the precise degree of carotid stenosis between 70% and 100%. Many studies and a recent review (1) conclude that collateral circulation is the major determinant of reserve/reactivity but provide little objective evidence for this. While published reserve or reactivity results support the hypothesis that patients with significant carotid stenosis and little or no reserve blood flow are at a significantly higher risk of stroke than those with “adequate” reserve, it is not possible to reach this conclusion in specific patients. A patient specific index that quantitates the value of reserve cerebral blood flow in predicting cerebral ischemia in patients with severe carotid stenosis is clearly needed.

A common assumption is that cerebrovascular reserve is normally a 2:1 proportion. This is correct if mean arterial pressure is near 100 mmHg and the lower cerebral perfusion threshold is 50mmHg. With no carotid stenosis the maximum Q/Qn ratio is 100/50 or 2:1. However, if Pa is 90mmHg the ratio is 1.8:1, or if Pa is 120mmHg the ratio is 2.4:1. With severe carotid stenosis reserve cerebral blood flow Qr is much less than the maximum with a ratio approaching 1:1.

Clinically measured ratios of reserve Qr/Qn and reactivity Vmax/Vn for patients considered to have adequate-normal reserve are 1.2 to 1.3 (1,2, 11). At 70.7% stenosis and 20% reserve and Pa = 120, 100, and 80 mmHg the Rw are 0.14, 1.0 and 1.3 respectively. This is a highly variant estimate of cerebral blood flow reserve. In contrast the zero reserve blood flow has close to acceptable Rw values at Pa = 100mmHg (Rw = 2.0) an excellent low Rw value at Pa = 80mmHg (Rw = 0.4).

Results at about 70% stenosis have little value. Repeating these model solutions at 81.4% carotid stenosis gives Rw = 3.0, 0.9 and 0.15 for normal 20% and ∞, ∞ and 1.5 for zero reserve. Percent reserve is clearly not an acceptable definition of adequate or impaired cerebral blood flow. The variance of Rw for both adequate and impaired is far too large, even at 80%. In contrast, treating measured cerebral blood flow reserve as a percentage of maximum reserve gives much tighter Rw variance. For example at 81.6% stenosis (Rc = 2.0), Rw = 0.6, 0.4 and 0.3 for 25% reserve (adequate) versus Rw = 18, 13 and 8 for zero reserve (impaired) respectively when Pa is 120, 100 and 80mmHg. Reserve cerebral blood flow is best predicted by the percent of maximum reserve blood flow not the percent of normal flow. This is how the Rw model solution treats reserve.

### Effect of Induced Cerebral Vasodilation on Collateral Circulation

The collateral circulation is primarily from the circle of Willis with three inflow/outflow vessels, the two internal carotid arteries and the Basilar artery. If induced cerebral vasodilation has a significant effect on these structures the model hemodynamic solutions will be adversely modified. Several studies aimed at this possibility failed to identify any significant change in the anatomy of circle of Willis (11) or acute changes in Willis pathways (12). In most patients the two carotid systems have good communication via the anterior communicating arteries (13,14). However in some patients the posterior inflow may be adequate to supply the hemispheres with bilateral severe carotid stenosis or stenosis/occlusion. Induced vasodilation of the cerebellum should not affect basilar artery blood flow potential. In contrast, vasodilation of the contralateral hemisphere will increase its blood flow with the potential to alter Willis hemodynamics. The presence of contralateral carotid stenosis inserts a resistance in series with the collateral circulation as seen by the ipsilateral vascular system (this model). This contralateral carotid resistance is determined by percent diameter stenosis, Rc2. This means that the patient’s collateral vascular resistance Rw is increased by Rc2. If the contralateral stenosis is mild, less than 50% diameter, Rc2 is less than o.32. For example, if an average patients Rw is 1.0 a mild contralateral stenosis will add 0.32 at most, or Rw = 1.32. In contrast, if the contralateral stenosis is severe, Rc2 is greater than 1.0 and the added resistance, Rw = 2.0 may have a significant adverse effect on blood flow reserve. This is more complicated, includes a potential steal or impaired collateral blood supply. Contralateral stenosis more than mild requires extensive model modification and is outside the scope of this study.

### Predictive Values of Patient Specific Collateral Vascular Resistance, Rw

Patient specific collateral vascular resistance predicts the adequacy of cerebral blood flow and blood flow reserve, to prevent ischemic symptoms or stroke with carotid stenosis progression. If cerebral perfusion pressure decreases to or falls below the lower threshold for brain auto regulation, (Pp = 50mHg used herein), three hemodynamic events occur. First, all reserve blood flow is utilized, second, regional cerebral blood flow is reduced below normal and third, cerebral blood flow is pressure dependent. At the intersection of cerebral blood flow auto regulation and pressure dependency Pp = 50mHg and the model equations is (Pa – 50) = RtQn. At carotid occlusion Rc = ∞ and Rt = Rw. Thus; Rw = Pa/Qn - 50/Qn and with normal cerebral blood flow 50/ml/min/100g, Rw = Pa/50 – 1.0. Therefore Rw = 0.6, and 1.4 when Pa = 80, 100, 120mmHg respectively. These are conservative values that predict normal cerebral blood flow, Qn, while exhausting blood flow reserve. When cerebral perfusion pressure, Pp drops below the lower auto-regulation threshold of 50mmHg the equation is Rw = Pa/Q - Pp/Q. In this pressure dependent model component Rb = 1.0 and Pp = Q. To maintain Q > 30ml/min/100g, the threshold for ischemic symptoms, Rw must be less than 1.0,1.4 and 1.8 for Pa of 80, 100 and 120mmHg respectfully. Similarly, to prevent cerebral infarction at Q=18 ml/min/100g, Rw must be less than 1.24, 1.64 and 2.04 respectively for Pa of 80, 100 and 120mmHg. In summary, for patients with relatively normal mean systemic arterial pressure of 80-120mmHg an Rw value less than 1.0 should be protective unless there is chronic low blood pressure. Approximately half of asymptomatic patients with significant carotid stenosis are in this group. The smaller the Rw value the greater protection. Conversely, patients with Re values greater than 1.0 are at increased risk while those with Rw > 3.0 should definitely be considered for carotid surgery. Collateral vascular resistance, Rw, is a unifying variable and index to predict the degree of cerebral vascular reserve.

## Discussion

The results of this study support the hypothesis that patients with significant ipsilateral carotid stenosis are dependent on collateral circulation to maintain adequate regional cerebral blood flow and blood flow reserve to prevent ischemic symptoms and/or stroke. There are three major findings.

1. The presence and amount of ipsilateral cerebrovascular reserve blood flow is best determined by patient specific numerical values of collateral vascular resistance, Rw. This hemodynamic index variable is a function of two independent variables, systemic arterial pressure and percent carotid stenosis.
2. The presence and amount of ipsilateral cerebrovascular reserve blood flow is almost entirely due to collateral circulation. Carotid artery blood flow reserve is completely utilized between 56% and 76% diameter stenosis dependent on mean systemic arterial pressure from 80mmHg to120mmHg. At mild to slightly moderate carotid stenosis the primary source of cerebrovascular reserve blood flow is the carotid artery.
3. Patient specific collateral vascular resistance, Rw, can be calculated from the model equations solution using measured values of percent diameter stenosis, mean systemic arterial pressure and reserve cerebral blood flow.

The clinical value of calculating a patient’s collateral vascular resistance value, Rw, is stroke risk stratification and advisability for carotid endarterectomy. The prospective randomized trials on asymptomatic patients with significant carotid stenosis were performed over two decades ago and prior to current best medical management with statins and anti-platelet drugs and improved carotid endarterectomy outcomes. It was found that it took 5-years to equate stroke prevention and risk of surgery. While part of this was due to poor surgical outcomes the reality is that many of these patients will not benefit from surgery. Modern current trials are likely to similarly indicate that surgical repair is not indicated. However, there remains a sub-set of asymptomatic patients with asymptomatic significant carotid stenosis that would benefit from carotid endarterectomy.

Criteria for selection should include an established perioperative stroke-mortality rate less than 2% by the surgeon/unit and the risk of cerebral ischemia if carotid stenosis progresses as predicted by the patient’s collateral vascular resistance value, Rw.

A meta-analysis of cerebrovascular reserve or reactivity in symptomatic and asymptomatic patients with greater than 70% carotid found that those with “normal” or “adequate” reserve-reactivity had a statistically significantly lower 3-year stroke rate than those patients with “inadequate” of absent reserve or reactivity (2). The criteria for “normal or adequate” and “inadequate” reserve or reactivity was not given. The individual publication results were assigned a binary value. This precludes any accurate estimate of the actual volumetric cerebral blood flow values. This, coupled with the failure to consideration the effect of systemic arterial pressure and precise percent carotid stenosis data on cerebral blood flow reserve negates using measured reserve results to predict patient specific collateral vascular resistance, Rw, values.

In theory, accurate values of three patient specific variables are necessary to calculate Rw. These are systemic arterial pressure, percent carotid stenosis and cerebral blood flow measured at normal and maximum brain vasodilation. While mean systemic arterial pressure can be easily determined, accurate and precise percent stenosis and cerebral blood flow are difficult to obtain. The standard non-invasive method of measuring percent diameter carotid stenosis is qualitatively based on various mid-stream Doppler velocities. Highly accurate NASCET percent diameters are not currently possible with ultrasound. The current best ultrasound estimate of significant carotid stenosis is moderate (50-69%) and severe (70-99%). This is partly due to the effect of collateral vascular resistance on carotid blood flow at high-grade stenosis, carotid velocity being directly related to flow (4). Of the three independent variables that determine patient specific collateral vascular resistance only accurate systemic arterial pressure is currently reliable. Future studies should strive to obtain accurate and precise percent stenosis based on anatomic area stenosis, (X^2^) and measurement of total or percent of maximum reserve blood flow.

Figures 1, 2, 3 and 4 illustrate the dependence of reserve and actual cerebral blood flow on systemic arterial pressure and degree of carotid stenosis as given by patient specific collateral vascular resistance curves, Rw. Figure 1 is maximum carotid artery blood flow over a range of mean systemic pressures when there is no collateral contribution. In the critical pressure dependent model component absence of collateral blood flow is predicted to result in profound cerebral ischemia with high-grade stenosis. This is theoretical, as a Ps value of zero has never been reported. In Figure 2, (Rw = 3.0), there is a large variance in reserve cerebral blood flow with systemic pressure at 70% stenosis. This is deceptive because at 80%-90% stenosis or occlusion the Rw curve predicts significantly impaired cerebral blood flow. Clearly selection of a safe Rw value is not easy. Figure 3 predicts that a patient with Rw values less then one (Rw < 1) tolerance of carotid occlusion. This represents approximately half of patients having carotid surgery as determined by carotid stump pressure measurements. Figure 4 (Rw = 0.5) basically confirms the predicted cerebral hemodynamic advantage of having an excellent circle of Willis. If only patient specific Rw vales were easily obtainable/measurable. Patients with Rw curves to the left on Figure 2, the upper 18% of patients with a Rw > 3.0, are clearly at risk for cerebral ischemia unless systemic arterial pressure is very high. Future clinical trials in asymptomatic patients with progressive carotid artery stenosis should include systemic arterial pressure and percent stenosis at the time of cerebral reserve or reactivity measurement, in addition to plaque stability analysis.

### Model Assumptions and Normal Values

The model requires three assumptions, a lower cerebral perfusion pressure threshold of auto-regulation value, a normal auto-regulated cerebral blood value and a functional relationship between carotid vascular resistance and the degree of carotid stenosis. The lower perfusion pressure threshold used, 50mmHg, was originally found by Lassen (3). He also measured the normal auto-regulated cerebral blood flow value used, Qn = 50ml/min/100g. These two normal values determine the minimum value of cerebral vascular resistance to be 1.0 (50mmHg/50 ml/min/100g = 1.0) at the junction between auto-regulation and pressure dependency. Later studies suggest that the lower perfusion pressure cut point and normal regional cerebral blood flow may be slightly higher, Pp = 60mmHg and Qn = 55 ml/min/100g. If these two values were used in the model the cerebral vascular resistance low value would be Rb = 60/55 = 1.09, not 1.0. This would change the model solutions slightly but the general relationships between the variables remain unchanged. Similarly, the maximum value of cerebral vascular resistance, Rb = Pa/Qn, (mean systemic pressure/normal auto-regulated flow) change only slightly. Carotid vascular resistance, Rc, is assumed to be equal to fractional percent diameter stenosis X where Rc = 1/[(1/X^2^ - 1)]. This energy consumption vascular stenosis model has been shown to correlate well with clinically measured values of critical carotid artery stenosis, predicting a cut-point between 60% and 70% diameter, as given in Figure 1. This assumption is key to the prediction that carotid artery blood flow reserve expires between 60% and 76% stenosis depending on the magnitude of mean arterial pressure (4). Not an assumption, but an established model solution with carotid occlusion, Rw = Rb(Pa/Ps - 1), is a patient specific index variable that is used to solve the hemodynamic model for cerebral blood flow.

### Other Findings of Interest

Cerebral blood flow and blood flow reserve as given by patient specific cerebral vascular resistance Rw curves are predicted to decrease rapidly between 70% to 90% stenosis, as highlighted in the Figures. The clinical utility of using blood flow solutions for predicting the risk of cerebral ischemia may increase with the degree of carotid stenosis. Selecting a safe Rw value at 70% stenosis based on reserve blood flow is difficult because of large reserve variability. Figures 2 and 7 illustrates this for patients with a mean systemic blood pressure of 100 mmHg or higher. Patient specific Rw values as high as 3.0 predict normal cerebral blood flow and good reserve at 70% stenosis but cerebral infarction with carotid occlusion. In Figure 7 the Rw = 5.7 curve has a 27ml/min/100g reserve at 70% stenosis but stroke at carotid occlusion. Blood flow predictions at 80% stenosis are much more reliable. Hypertension may increase reserve blood flow but it also associated with higher Rw values and a more severe reduction of reserve and actual cerebral blood flow at higher degrees of stenosis.

Patients with cerebral aneurisms undergoing staged acute internal carotid artery occlusion have a 12.5% cerebral infarction rate (15). This is consistent with what might be expected in patients with the low cerebral blood flows less than 25ml/min/100g at 100% stenosis. Figure 2 indicates that approximately 16% of 1,360 patients with significant carotid stenosis having carotid endarterectomy (4, 6) have collateral vascular resistance values higher than 3.0 and cerebral blood flows blood flows less than 30ml/min/100g with carotid occlusion. Conversely Figures 3 and 4 indicate that approximately half of that same 1,360 patient cohort having carotid surgery with collateral vascular resistance of 1.0 or less have normal cerebral blood flow and most have vascular reserve with carotid occlusion. Accordingly, this latter group of patients may have a low risk of stroke with carotid progression and should not be considered for carotid endarterectomy.

### Model Limitations

This is a two component, one-dimensional, lumped parameter, steady flow model based on the principle of energy conservation. All models of the cerebrovascular system that includes carotid artery stenosis must address and account for carotid stenosis turbulence. While the turbulent carotid blood flow problem suggests that little might be expected from analysis of such a simple biomechanical system this is not the case. Three assumptions are sufficient to solve the model equations for cerebral blood flows. They are steady blood flow, a simple arterial stenosis energy dissipation constitutive equation and a previously established model solution for collateral vascular resistance. The assumption of steady, not physiologic pulsatile, blood flow is a major limitation. A further limitation is neglecting the small viscous laminar Poiseuille flow energy losses. It may be a leap of faith to assume that valid pulsatile flow solutions to an energy equation integrated over a cardiac cycle reduce to the results of this model. However the results using steady flow and the turbulent flow energy loss constitutive equation may justify the means. This is made possible by assuming that carotid stenosis vascular resistance, Rc, is proportional to percent area stenosis X^2^ and Rc = 1/[(1/X^2^) – 1] where X is fractional percent carotid stenosis. This is similar to proposed Bernoulli turbulent energy losses that equal the decrease in kinetic energy. This simple second power function correlates well with measured values of critical carotid stenosis (4).

As mentioned above the small laminar flow viscous pressure gradients are neglected. Similarly, the 5 to 6 mmHg cerebral venous/ cerebral fluid pressure is not considered. This is equivalent to treating mean systemic pressure as normalized, actual arterial pressure minus the venous pressure at the same elevation.

### Conclusions

The value of hemodynamic models is not to support existing knowledge or theories but rather to aid in conceptualizing and understanding complex relationships between variables. In this study the model solutions for actual and reserve cerebral blood flow in patients with significant carotid artery stenosis predict that reserve cerebral blood flow is significantly determined by two patient specific independent variables, systemic arterial pressure and percent carotid stenosis. Reserve blood flows are extremely variable when diameter carotid stenosis is less than 75% to 80%. At higher percent carotid stenosis the majority of actual cerebral blood flow and all reserve blood flow is predicted to be collateral in origin. The unifying variable that best predicts cerebral blood flow is collateral vascular resistance, Rw. It is patient specific and dependent on both systemic arterial pressure and precise degree of carotid stenosis. Approximately half of patient with asymptomatic severe carotid stenosis are predicted to have collateral vascular resistance values less than 1.0, Rw < 1.0, maintaining adequate cerebral blood flow if carotid stenosis progresses to occlusion. Conversely, patients with Rw > 1.0 may benefit from carotid surgery to prevent cerebral ischemia with stenosis progression. Collateral vascular resistance can be calculated from the model solution by inserting each patient’s accurately measured systemic pressure; percent carotid stenosis and total cerebral blood flow during induced maximal cerebral vasodilation.

## Data Availability

All clinical data used is published and referenced.

## Notes

### Competing Interest Statement

The authors have declared no competing interest.

### Funding Statement

none

## References

1. Aliev VA, Semenyutin VB, Panuntser GK. Predictive value of cerebrovascular reserve in patients with carotid artery stenosis for choosing treatment strategy. International J Pathology and Clinical Research 2019;DOI:10.23937/2469-5807/1510086.

2. Gupta A, Chazen JL, Hartman M, Delgado D, Anumula N, Shao H et al. Cerebrovascular reserve and stroke risk in patients with carotid stenosis or occlusion: A systemic review and meta-analysis Stroke 2012;43: 2884–91.

3. Lassen NA Cerebral blood flow and oxygen consumption in man. Physiol Rev 1959;39: 183–238.

4. Archie JP. A hemodynamic model to predict regional cerebral blood flow and blood flow reserve in patients with carotid stenosis. Med Rx IV 2020. 07.21.20158840.

5. Powers WJ. Cerebral hemodynamics in ischemic cerebrovascular disease. Ann Neurol 1991;29: 231–40.

6. Archie, JP. A fifteen-year experience with carotid endarterectomy after a formal operative protocol requiring highly frequent patch angioplasty. J Vasc Surg 2000: 31: 24–35.

7. Vagal JL, Leach M, Ferandez-Ulloa M, Zuccarello M. The acetazolamide challenge: Techniques and applications in the evaluation of chronic cerebral ischemia. Am J Neurol 2009;30: 376–84.

8. Dogal A, Lam AM. Cerebral auto-regulation and anesthesia. Curent Opinion in Anesthesiology 2009, 22:547–52.

9. Boysen G, Engell HC, Pistolese GR, Fiorani P, Angoli A, Lassen NA. On the critical lower level of cerebral blood flow in man with particular reference to carotid surgery. Circulation 1974;49: 1023–5.

10. Dahl A, Lindegaard KF, Russell D, Nyberg-Hansen R, Rootwelt K, Sorteberg W, Nires H. A comparison of transcranial Doppler and cerebral blood flow studies assass cerebral vasoreactivity. Stroke 1992;23:15–9.

11. Stojanovic N, Stefanoric I, Kostic A. Radiasarejevic M, Stejanor S, Petrovic S. Analysis of the symetric configurations of the Circle of Willis in a series of autopsied cases. Vojnosanit Preyl 2015; 74: 356–60.

12. Zhu G, Yuan Q, Yang J, Yeo JH. The role of the circle of Willis in internal carotid artery stenosis and anatomical variations: a computational study based on a patient specific three-dimensional model. BioMedical Engineering On Line 2015;14:107–36.

13. Vernieri F, Pasqualetti P, Matteis M, Passayelli F, Troisi E., et.al. Effect of collateral flow on cerebral vasomotor reactivity on outcome of carotid artery occlusion. Stroke 2001;32: 1552–8.

14. Reinhard M, Roth M. Muller T, Timer J et.al. Cerebral auto regulation in carotid artery occlusive disease assessed from spontaneous blood pressure fluctuations by the correlation coefficient index. Stroke 2003;34:2138–44.

15. Linskey WE, Jungreis CA, Yonas H, Hirch WL, Sekhar LN,Horton JA, et al. Stroke risk after abrupt internal carotid artery sacrifice: Accuracy of preoperative assessment with balloon test occlusion and stable Xenon-enhanced CT. Am J Neuroradiol 1994;15:829–43.

